# Characteristics, outcome and predictors of in-hospital mortality in an elderly population from a SARS-CoV-2 outbreak in a long-term care facility

**DOI:** 10.1101/2020.06.30.20143701

**Authors:** Enrico Maria Trecarichi, Maria Mazzitelli, Francesca Serapide, Maria Chiara Pelle, Bruno Tassone, Eugenio Arrighi, Graziella Perri, Paolo Fusco, Vincenzo Scaglione, Chiara Davoli, Rosaria Lionello, Valentina La Gamba, Giuseppina Marrazzo, Maria Teresa Busceti, Amerigo Giudice, Marco Ricchio, Anna Cancelliere, Elena Lio, Giada Procopio, Francesco Saverio Costanzo, Daniela Patrizia Foti, Giovanni Matera, Carlo Torti, IDTM UMG COVID-19 Group

## Abstract

Since December 2019, coronavirus disease 2019 (COVID-19) pandemic has spread from China all over the world, many COVID-19 outbreaks have been reported in long-term care facilities (LCTF). However, data on clinical characteristics and prognostic factors in such settings are scarce. We conducted a retrospective, observational cohort study to assess clinical characteristics and baseline predictors of mortality of COVID-19 patients hospitalized after an outbreak of SARS-CoV-2 infection in a LTCF.

A total of 50 patients were included. Mean age was 80 years (SD, 12 years), and 24/50 (57.1%) patients were males. A total of 42/50 (84%) patients experienced symptoms of SARS-CoV-2 infection. The overall in-hospital mortality rate was 32%. In Cox regression, significant predictors of in-hospital mortality were: hypernatremia (HR 9.12), lymphocyte count <1000 cells/µL (HR 7.45), cardiovascular diseases other than hypertension (HR 6.41), and higher levels of serum interleukin-6 (IL-6, pg/mL) (HR 1.005).

Our study shows a high in-hospital mortality rate in a cohort of elderly patients with COVID-19 and hypernatremia, lymphopenia, CVD other than hypertension, and higher IL-6 serum levels were identified as independent predictors of in-hospital mortality. Further studies are necessary to better understand and confirm our findings in the setting of a LTCF outbreak of COVID-19.

## Introduction

Since SARS-CoV-2 pandemic has spread from China all over the world, more than 3 700 000 people have been infected, with almost 260 000 of reported deaths until May 8^th^, 2020 [1]. The spectrum of clinical manifestations of SARS-CoV-2 associated disease (COVID-19), varies from the absence of symptoms to severe disease, eventually leading to death. The actual prevalence of asymptomatic patients remain unknown, although it has been reported as high as 50% [2]. Among symptomatic patients, COVID-19 has been reported to be severe and critical in 14% and 5% patients, respectively [3]. Since the first clinical reports on COVID-19, older age and coexisting comorbidities, in particular cardiovascular or cerebrovascular diseases, have been highlighted as risk factors associated with an adverse outcome of COVID-19 [4-7]. These clinical characteristics are common among residents in Long-Term Care Facility (LTCFs). Noteworthy, these institutions can act as incubators of infection and elderly who live in LTCF are at higher risk of SARS-CoV-2 infection [8]. Several COVID-19 outbreaks have been reported in different countries, including Italy, with high mortality rates, reaching 49-64% in some countries [9]. Although there are studies describing characteristics of such patients and dynamics of transmission in this setting [10], there is a lack of analyses investigating predictors of mortality in geriatric patients. These results are important to guide clinical management. Herein, we report clinical characteristics, outcome (in-hospital mortality), and prognostic factors in a cohort of 50 patients as part of an outbreak of SARS-CoV-2 infection in an Italian LTCF.

## Results

In the present study, clinical characteristics and outcome of a cohort of 50 patients diagnosed with COVID-19 who were transferred from a LTCF to the Infectious and Tropical Disease Unit of “Mater Domini” Teaching Hospital, Catanzaro, Italy are described. Out of 50 patients, 49 were diagnosed with COVID-19 by positive SARS-CoV-2 molecular test conducted on nasopharyngeal swab; the remaining patient was diagnosed on the basis of epidemiological link and clinical/radiological findings. Mean age was 80 years (SD, 12 years), and 24/50 (57.1%) patients were males. A total of 42/50 (84%) patients experienced COVID-19 symptoms with a mean time between onset of symptoms and hospitalization of 6 days (SD, 3 days), whereas the remaining 8 (16%) patients remained asymptomatic.

In **Table 1**, clinical and demographic characteristics and outcome of the study patients are reported overall and according to presence or absence of clinical symptoms. The majority of patients suffered from cardiovascular diseases (CVD, 82%) and/or neurological diseases (52%); 52% of patients presented more than two comorbidities.

**Table 1.** Clinical and demographic characteristics and outcome of patients with COVID-19 coming from a LTCF, according to presence of clinical symptoms.

| Variables | Total<br>(n=50) | Symptomatic<br>(n = 42) | Asymptomatic<br>(n = 8) | P values |
| --- | --- | --- | --- | --- |
| *Demographic information* |  |  |  |  |
| Male sex | 28 (57.1) | 24 (57.1) | 4 (50) | 0.70 |
| Age (years, mean ± SD) | 80 ± 12 | 79 ± 12 | 86 ± 8 | 0.09 |
| *Comorbidities and previous drug therapies* |  |  |  |  |
| Cancer | 9 (18) | 8 (19.1) | 1 (12.5) | 0.65 |
| Cardiovascular diseases (CVD) | 41 (82) | 34 (80.9) | 7 (87.5) | 0.65 |
| CVD other than hypertension | 19 (38) | 16 (38.1) | 3 (37.5) | 0.97 |
| Psychiatric disorders | 15 (30) | 12 (28.6) | 3 (37.5) | 0.61 |
| Neurologic diseases | 26 (52) | 23 (54.7) | 3 (37.5) | 0.37 |
| Obesity | 5 (10) | 5 (11.9) | 0 | 0.30 |
| Diabetes mellitus | 11 (22) | 11 (26.2) | 0 | 0.10 |
| COPD | 9 (18) | 9 (21.4) | 0 | 0.14 |
| Chronic renal failure | 20 (40) | 18 (42.8) | 2 (25) | 0.34 |
| >2 coexisting comorbidities | 26 (52) | 25 (59.5) | 1 (12.5) | **0.01** |
| Polipharmacy | 36 (72) | 29 (69.1) | 7 (87.5) | 0.28 |
| Number of medications (mean ± SD) | 6 ± 3 | 6 ± 3 | 7 ± 3 | 0.54 |
| ACE-I/ARB therapy | 16 (32) | 12 (28.6) | 4 (50) | 0.23 |
| *Outcome scores and mortality* |  |  |  |  |
| *Brixia* score (mean ± SD) | 7 ± 3 | 7 ± 3 | 7 ± 3 | 0.96 |
| CALL score (mean ± SD) | 11 ± 2 | 11 ± 2 | 11 ± 1 | 0.43 |
| In-hospital mortality | 16 (32) | 14 (33.3) | 2 (25) | 0.64 |

Abbreviations: CVD, cardiovascular diseases; ACE-I, angiotensin-converting-enzyme inhibitors; ARBs, angiotensin-receptor blockers.

There were no statistical differences between symptomatic and asymptomatic patients in terms of demographic characteristics and specific comorbidities; only the coexistence of more than two comorbidities was significantly more frequent in patients who displayed symptoms compared to the asymptomatic ones (59.5% vs. 12.5%, P=0.01). The overall in-hospital mortality rate was 32% (16/50) and did not differ significantly between symptomatic and asymptomatic patients (33.3% vs. 25%, P=0.64).

A total of 38/50 (76%) patients received combination therapy with hydroxychloroquine plus azithromycin according to the study protocol by Gautret et al.[14], with frequent electrocardiographic monitoring and without cardiac complications. Moreover, all the patients received anticoagulant therapy with enoxaparin (44 patients) or fondaparinux (6 patients); in 26/50 patients (52%) anticoagulants were administered at therapeutic dosage, whereas in the remaining 24 (48%) at prophylactic dosage. Finally, on the basis of clinical judgment, in 25/50 (50%) patients corticosteroid therapy with intravenous methylprednisolone was administered. Two patients were treated with tocilizumab single subcutaneous 162 mg administration, as described elsewhere[15].

### Risk factors for in-hospital mortality

Analyses of risk factors for in-hospital mortality have been conducted in 48/50 patients; two patients were excluded because they died few hours from hospital admission and thereof complete dataset was lacking. At univariate analysis (described in **Table 2**), variables associated with in-hospital mortality were: CVD other than hypertension (P<0.001), presence of dyspnea at hospital admission (P<0.001), higher *Brixia* and CALL scores (P= 0.01 and 0.003, respectively), higher levels of gamma glutamyl transferase (P=0.002), a blood sodium level >145 mmol/L (P=0.001), lymphocyte count <1000 cells/µL (P=0.02), higher levels of D-dimer (P=0.002) and serum Interleukin-6 (P=0.007), C reactive protein > 50 mg/L (P=0.01), and corticosteroid therapy (P<0.001); psychiatric disorders were positively associated with survival (P=0.03).

**Table 2.** Univariate analysis of risk factors for mortality of 48 patients with COVID-19 coming from a LTCF.

| Variables | Non-survivors<br>(n = 14) | Survivors<br>(n = 34) | OR (95% IC) | P values |
| --- | --- | --- | --- | --- |
| *Demographic information* |  |  |  |  |
| Male sex | 5 (35.7) | 21 (61.7) | 0.34 (0.07-1.47) | 0.09 |
| Age (years, mean ± SD) | 85 ± 8 | 78 ± 13 | - | 0.06 |
| *Comorbidities* |  |  |  |  |
| Cancer | 4 (28.6) | 4 (11.7) | 3.00 (0.45-19.01) | 0.15 |
| Cardiovascular diseases (CVD) | 12 (85.7) | 28 (82.3) | 1.28 (0.19-14.73) | 0.77 |
| CVD other than hypertension | 11 (78.8) | 7 (20.6) | 14.14 (2.58-93.97) | **<0.001** |
| Psychiatric disorders | 1 (7.1) | 13 (38.2) | 0.12 (0.01-1.05) | **0.03** |
| Neurologic diseases | 9 (64.3) | 17 (50) | 1.80 (0.45-8.26) | 0.36 |
| Obesity | 1 (7.1) | 4 (11.7) | 0.57 (0.01-6.68) | 0.63 |
| Diabetes mellitus | 3 (21.4) | 8 (23.5) | 0.88 (0.12-4.68) | 0.87 |
| COPD | 4 (28.6) | 4 (11.7) | 3.00 (0.45-19.01) | 0.15 |
| Chronic renal failure | 5 (35.7) | 14 (41.2) | 0.79 (0.17-3.37) | 0.72 |
| >2 comorbidities | 7 (50) | 18 (52.9) | 0.88 (0.21-3.71) | 0.85 |
| *Symptoms (Total)* | 12 (85.7) | 28 (82.3) | 1.28 (0.19-14.73) | 0.77 |
| Days from symptoms to hospitalization (days, mean ± SD) | 5 ± 3 | 6 ± 3 | - | 0.62 |
| Fever | 10 (71.4) | 23 (67.6) | 1.19 (0.26-6.39) | 0.79 |
| Cough | 3 (21.4)) | 14 (41.2) | 0.38 (0.06-1.89) | 0.19 |
| Dyspnea | 5 (35.7) | 0 | - | **<0.001** |
| *Polipharmacy* | 11 (78.6) | 25 (73.5) | 1.32 (0.25-8.98) | 0.71 |
| Number of medications (mean ± SD) | 6 ± 3 | 6 ± 3 | - | 0.70 |
| ACE/ARB | 3 (21.4) | 13 (38.2) | 0.44 (0.06-2.15) | 0.26 |
| *Brixia* score (mean ± SD) | 9 ± 2 | 7 ± 3 | - | **0.01** |
| CALL core | 12 ± 1 | 11 ± 1 | - | **0.003** |
| Temperature ≥ 37.5°C | 3 (21.4) | 4 (11.7) | 2.04 (0.25-14.11) | 0.38 |
| Heart rate ≥ 85 beats per minute | 6 (42.8) | 10 (29.4) | 1.8 (0.39-7.77) | 0.36 |
| Median blood pressure (mmHg, mean ± SD) | 92 ± 17 | 93 ± 14 | - | 0.84 |
| Oxygen saturation ≤ 94% | 7 (50) | 8 (23.5) | 3.25 (0.71-14.62) | 0.07 |
| Glycemia (mg/dl, mean ± SD) | 139 ± 72 | 121 ± 71 | - | 0.43 |
| Creatinine ≥ 1.20 mg/dl | 4 (28.6) | 11 (32.3) | 0.83 (0.15-3.81) | 0.79 |
| Aspartate transaminase (UI/L, mean ± SD) | 132 ± 313 | 32 ± 14 | - | 0.06 |
| Gamma glutamyl transferase (UI/L, mean ± SD) | 99 ± 122 | 30 ± 22 | - | **0.002** |
| Blood sodium level > 145 mmol/l | 9 (64.3) | 6 (17.6) | 8.4 (1.69-43.31) | **0.001** |
| Ferritin level (ng/ml, mean ± SD) | 732 ± 571 | 464 ± 336 | - | 0.08 |
| Lactate dehydrogenase > 500 UI/L | 10 (76.9) | 19 (55.9) | 2.63 (0.53-17.19) | 0.18 |
| Lymphocytes count <1000 cells/μL | 10 (71.4) | 12 (35.3) | 4.58 (1.01-23.73) | **0.02** |
| Platlets count (cells/μl, mean ± SD) | 195 ± 88 | 197 ± 72 | - | 0.93 |
| D-dimer (mg/L, mean ± SD) | 5.41 ± 7.49 | 1.21 ± 1.05 | - | **0.002** |
| C reactive protein > 50 mg/L | 10 (71.4) | 11 (32.2) | 5.22 (1.13-27.20) | **0.01** |
| Interleukin-6 (pg/mL, mean ± SD) | 125 ± 189 | 34 ± 22 | - | **0.007** |
| Enoxaparin (therapeutic dose) | 9 (64.3) | 17 (50) | 1.8 (0.42-8.26) | 0.36 |
| Hydroxychloroquine plus azytromicin therapy | 9 (64.3) | 28 (82.3) | 0.38 (0.07-2.04) | 0.17 |
| Corticosteroid therapy | 13 (92.8) | 12 (35.3) | 23.83 (2.75-1057.36) | **<0.001** |

Abbreviations: CVD, cardiovascular diseases; ACE-I, angiotensin-converting-enzyme inhibitors; ARBs, angiotensin-receptor blockers.

At multivariable Cox regression, significant predictors of in-hospital mortality were: blood sodium level >145 mmol/L (HR 9.12, 95% CI 2.15-38.52; P=0.003), lymphocyte count <1000 cells/µL (HR 7.45, 95% CI 1.81-30.68; P=0.005), CVD other than hypertension (HR 6.41, 95% CI 1.51-27.22; P=0.01), and higher IL-6 serum levels (pg/mL) (HR 1.005, 95% CI 1.001-1.009; P=0.007) (**Table 3**).

**Table 3.**
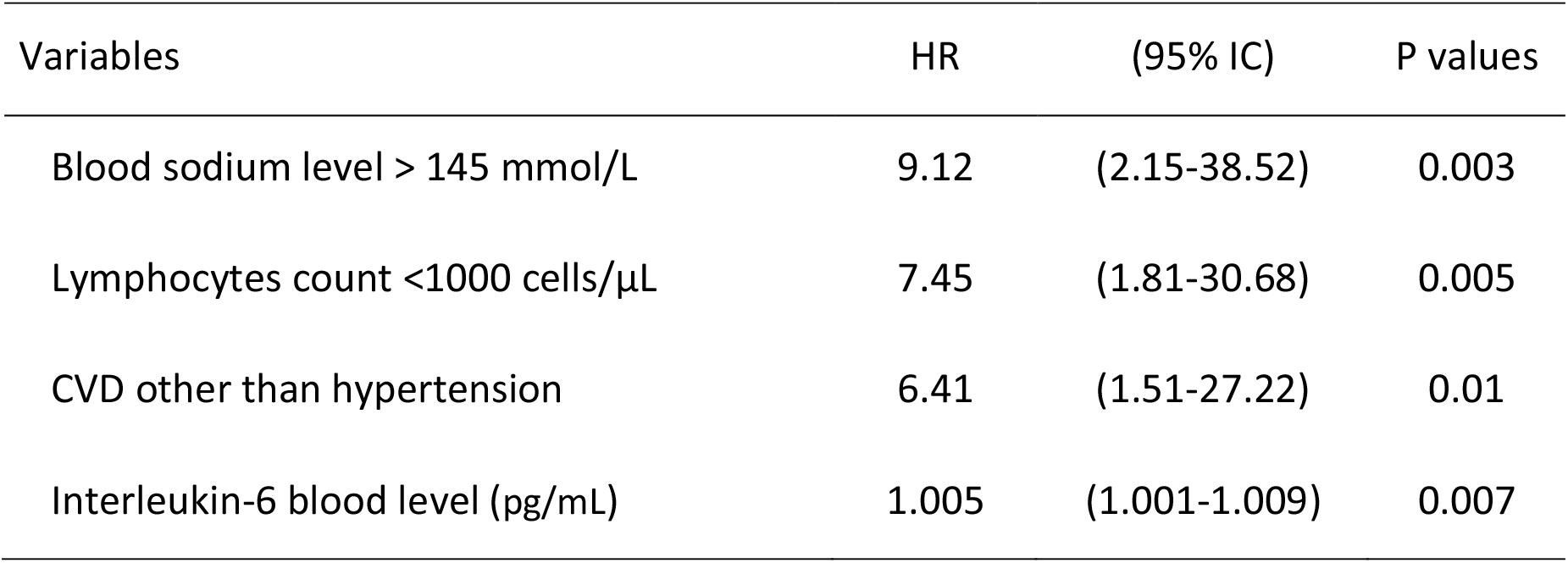
Cox regression analysis for in-hospital mortality in COVID-19 patients coming from a LTCF.

| Variables | HR | (95% IC) | P values |
| --- | --- | --- | --- |
| Blood sodium level > 145 mmol/L | 9.12 | (2.15-38.52) | 0.003 |
| Lymphocytes count <1000 cells/μL | 7.45 | (1.81-30.68) | 0.005 |
| CVD other than hypertension | 6.41 | (1.51-27.22) | 0.01 |
| Interleukin-6 blood level (pg/mL) | 1.005 | (1.001-1.009) | 0.007 |

## Discussion

In the present study, we found that in-hospital mortality among elderly resident in a LTCF and diagnosed with COVID-19 was 32%. This finding is in line with (or slightly lower than) preliminary data estimated in LTCFs in Italy, where mortality has been reported as high as 37.4%, much lower than reported in other countries (49-64%)[9,16]. It has however to be recognised that we report herein the in-hospital mortality, not taking into account possible deaths occurred inside the LTCF. Notwithstanding this consideration, taking into account mean age of our patients, i.e. 80 years, in-hospital mortality rate in our cohort is in line with the case fatality rate (30.4%) among patients with age ranging from 80 and 89 years in the general population as reported by the Italian National Institutes of Health (Istituto Superiore di Sanità)[17]. With regard to presence of clinical symptoms, a significant percentage of our patients (84%) was symptomatic at hospital admission; this percentage is higher compared to that reported in a cohort of residents of a LTCF during a COVID-19 outbreak in USA, in which only 13/23 (56%) of patients tested positive for SARS-CoV-2 presented clinical symptoms of COVID-19 [18]. In two serial point-prevalence surveys conducted during a SARS-CoV-2 outbreak in a Skilled Nursing Facility, on a total of 48 residents who tested positive, 27 (56%) were asymptomatic at the time of the first survey but, of these, 24 subsequently developed symptoms so that the definitive rate of symptomatic patients was 93.7%, slightly higher compared to that in our cohort [10]. Of note, we did not find any significant difference for in-hospital mortality between symptomatic and asymptomatic patients in our cohort. So it is possible that in elderly patients with multiple comorbidities, death occurs even without symptoms.

Multivariable Cox regression analysis showed that blood sodium level >145 mmol/L, lymphocyte count <1000 cells/µL, CVD other than hypertension, and higher IL-6 serum levels were independent predictors of in-hospital mortality.

In general, hypernatremia (defined as a sodium value higher than 145 mEq/L) is a condition frequently observed in patients (especially elderly) at the time of hospitalization and affects up to 9% of critical patients. Moreover, it has been reported that patients with hypernatremia have a significantly higher mortality rate compared to patients with normal values [19,20]. To the best of our knowledge, hypernatremia has been never recognized as independent risk factor for mortality in COVID-19 patients. Instead, of note, hypernatremia has been reported as a factor significantly associated with mortality in a small (n=25) cohort of patients suffering from severe acute respiratory syndrome (SARS) and ARDS [21]. Hypernatremia has been identified as a potential surrogate marker of sepsis, especially in elderly because a severe systemic infection can lead to a significant free water deficit and subsequent hypernatremia [22]. In addition, some evidence also suggests that sodium could represent an important promoter of immune response, improving the function of macrophages and T-lymphocytes [23]. Although our finding should be confirmed by further studies, it could be assumed that hypernatremia may have a role as a marker related to the severity of the inflammatory state in COVID-19 elderly patients.

Lymphopenia and cardiovascular diseases have been widely described as factors associated with a worse outcome in cohorts of COVID-19 patients [4-7,24]. Our study confirms the important clinical impact of these factors also in elderly patients. Of note, in line other studies, in our patients there was not any difference for in-hospital mortality between patients who were receiving angiotensin-converting–enzyme (ACE) inhibitors or angiotensin-receptor blockers (ARBs) and those not treated with these drugs [4].

The last independent predictor of mortality in our cohort was higher IL-6 serum levels. A cytokine storm, also known as cytokine release syndrome, has been suggested to be a major mechanism causing the more severe and often fatal clinical complications of COVID-19, such as ARDS and multi-organ dysfunction [25]. In particular, IL-6, which has been implicated in many immunological functions (e.g. B-cell stimulation and induction the production of acute phase proteins), can also activate the clotting pathway, vascular endothelial cells, and lead to a cytokine storm [26]. Gao et al. demonstrated that high levels of IL-6 and D-dimer were closely related to the occurrence of severe COVID-19 in adult patients [27]. Moreover, Wan et al. demonstrated that IL-6 serum levels can predict transition from mild to severe infection [28], and Liu et al. found that in Cox proportional hazard model IL-6 was an independent factor predicting severity of COVID-19 with a significant level >32.1 pg/mL [29]. In line with these findings, we have demonstrated that higher IL-6 serum levels can independently predict the risk of in-hospital mortality in a cohort of COVID-19 elderly patients.

This study has major limitations, including its retrospective nature and the small population size. By contrast, the main strength is that patients were all infected in a limited period of time, so this is an incident cohort, and analysis of outcome predictors could be more reliable.

To the best of our knowledge, the present is the first study investigating clinical characteristics, mortality and prognostic factors of a complete cohort of patients during a SARS-CoV-2 outbreak in a LTCF. We found that in-hospital mortality was 32%, in line with fatality rates reported in elderly patients in Italy. Importantly, hypernatremia, lymphopenia, CVD other than hypertension, and higher IL-6 serum level appeared to be independent predictors of in-hospital mortality. Further larger studies are necessary to better understand and confirm our findings, in order to rapidly identify characteristics associated with an adverse outcome among elderly suffering from COVID-19 and provide a more aggressive monitoring and care.

## Materials and Methods

We conducted a retrospective, single-centre, observational cohort study in order to describe clinical characteristics and baseline predictors of mortality of COVID-19 in hospitalized patients as part of an outbreak of SARS-CoV-2 infection in a LTCF.

Between March 27, 2020 and May 6, 2020, a total of 50 consecutive patients diagnosed with SARS-CoV-2 infection were transferred from a LTCF and admitted at Infectious and Tropical Disease Unit of “Mater Domini” Teaching Hospital, Catanzaro, Italy. The diagnosis of SARS-CoV-2 infection was established according to the WHO recommendations [11].

This study was notified to the Ethics Committee of the Calabria Region on May 13^th^, 2020 and conducted in in accordance with the declaration of Helsinki. The study was carried out using retrospectively collected and anonymized data. In Italy, such studies do not require ethical approval by an Ethics Committee as determined by the Italian Drug Agency note 20 March 2008 (GU Serie Generale n° 76 31/3/2008). The need for written informed consent was waived for patients owing to the retrospective nature of the study.

### Data collection

Data collected from the hospital charts and the laboratory database included patient demographics, underlying diseases, previous drug therapies (including angiotensin-converting– enzyme [ACE] inhibitors and angiotensin-receptor blockers [ARBs]), reported COVID-19 symptoms (i.e., fever, cough, dyspnea), clinical signs at hospital admission (i.e. body temperature, heart rate, median blood pressure, oxygen saturation level), laboratory findings at hospital admission (including serum interleukin-6 [IL-6], D-dimer, C reactive protein). All therapies administered for treatment of COVID-19 were also recorded. Chest X-ray (CXR) findings were classified according to the experimental CXR scoring system (*Brixia* score) validated by Borghesi et al. in hospitalized patients with COVID-19 pneumonia in order to quantify and monitor the severity and progression of COVID-19 [12]. The CALL score was also calculated in order to predict the progression of COVID-19 [13].

All information were entered in case report forms and then recorded in a specific database. Two researchers independently reviewed the data collection forms to double check the collected data. The outcome measured was in-hospital mortality during the study period. Survivor and non-survivor subgroups were compared in order to identify predictors of mortality.

### Statistical analysis

Continuous variables were compared with Student’s *t* test for normally distributed variables and the Mann-Whitney U test for non-normally distributed variables. Categorical variables were evaluated using the χ^2^ or two-tailed Fisher’s exact test. Odds ratios (ORs) and 95% confidence intervals (CIs) were calculated to evaluate the strength of any association that emerged. Values are expressed as means ± standard deviation (SD) (continuous variables) or as percentages of the group from which they were derived (categorical variables). Two-tailed tests were used to determine statistical significance; a P value of <0.05 was considered significant. Cox regression analysis was conducted to identify independent risk factors for 21-day mortality. Variables emerging from univariate analysis with P values of <0.05 were included in the Cox regression model. All statistical analyses were performed using the Intercooled Stata program, version 16, for Windows (Stata Corporation, College Station, Texas, USA). Description of statistical analysis was partially reported elsewhere [30].

## Data Availability

Data cannot be made publicly available due to ethical restrictions imposed by Italian legislation and our Ethics Committee

## Acknowledgements

We want to thank all our patients and our nurses. We also thank the Infectious Diseases and Tropical Medicine (IDTM) of the University “*Magna Graecia*” (UMG) COVID-19 Group, which is composed, besides the main authors, by the following: Domenico Laganà, Maria Petullà, Bernardo Bertucci, Angela Quirino, Giorgio Settimo Barreca, Aida Giancotti, Luigia Gallo, Angelo Lamberti, Maria Carla Liberto, Nadia Marascio, Adele Emanuela De Francesco.

## Author contributions statement

All authors had full access to all the data in the study and take responsibility for the integrity of the data and the accuracy of the data analysis. EMT and CT were responsible for study concept and design. FS, MCP, BT, EA, GP, PF, VS, CD, RL, VLG, GM, MTB, AG, MR, AC, EL, GP, FSC, DPF, GM were responsible for the acquisition, analysis, and/or interpretation of clinical or laboratory data. EMT, CT and MM were responsible for drafting the manuscript. EMT was responsible for statistical analysis. All authors subsequently revised the manuscript.

